# Increased uptake of tuberculosis preventive therapy (TPT) among people living with HIV following the 100-days accelerated campaign: A retrospective review of routinely collected data at six urban public health facilities in Uganda

**DOI:** 10.1101/2022.05.12.22274996

**Authors:** Joseph Musaazi, Christine Sekaggya-Wiltshire, Stephen Okoboi, Stella Zawedde-Muyanja, Mbazi Senkoro, Nelson Kalema, Paul Kavuma, Proscovia M. Namuwenge, Yukari C Manabe, Barbara Castelnuovo, Agnes Kiragga

**Author notes:** **Corresponding author:** (JM). **Author contributions:** JM, CSW, AK, SO, SZM, NK, and BC designed the assessment. JM led the training of the data collection teams. JM supervised and oversaw the data collection. JM analyzed the data. All co-authors contributed to the critical interpretation of the results. JM led the writing of the manuscript with substantial contributions from all co-authors. All co-authors reviewed and approved the final version of the manuscript.

## Abstract

**Introduction:** Tuberculosis preventive therapy (TPT) effectively decreases rates of tuberculosis reactivation in people living with HIV (PLHIV) who are at increased risk. The Uganda Ministry of Health launched a 100-day campaign to scale-up TPT in PLHIV in July 2019. We sought to examine the effect of the campaign on trends of TPT uptake and characteristics associated with TPT uptake and TPT completion among persons in HIV care. We retrospectively reviewed routinely collected data from 2016 to 2019 at six urban public health facilities in Uganda.

**Methods:** A cross-sectional review of the HIV care database and paper-based TPT registers at six public health facilities in Kampala, Uganda. Estimated trends of prevalence of TPT (given as Isoniazid preventive therapy [IPT]) uptake and treatment completion across the 4 years, among PLHIV aged 15 years and above, and factors associated, were examined using Poisson regression model with robust standard errors using generalized estimating equation (GEE) models.

**Results:** On average a total of 43,215 patients aged 15 years and above were eligible for TPT each calendar year at the six health facilities. More than 70% were females and median age was 34 years (inter-quartile range 28 to 41 years on overall). There was consistently low TPT uptake from 2016 to 2018, but as expected, the uptake significantly increased by about 25% (relative increase) from 2.5% of eligible PLHIV in 2018 to 64.8% in 2019 (prevalence of TPT uptake 4.5% (1,746/38,704), 4.4% (1,743/39,630), 2.5% (1,018/40,390), and 64.8% (26,478/40,867) during 2016, 2017, 2018 and 2019 respectively). TPT uptake in 2019 was 26.0 times higher compared to that in 2018 (adjusted prevalence ratio [aPR] = 26.0 [95%CI 24.5, 27.7], P-value<0.001). Also, TPT completion was consistently high at about 80% between years 2016 – 2018 but dramatically increased by 17% (relative increase) in 2019 compared to that in 2018 (prevalence of TPT completion was 81.2%, 76.3%, 82.6% and 96.5% in year 2016, 2017, 2018 and 2019 respectively). The increase in TPT completion prevalence from 2018 to 2019 remained significant even after adjusting for patients’ baseline characteristics (aPR [95%CI] = 1.12 [1.04, 1.21], P value=0.003). Not on ART or newly started on ART compared ART experienced were associated with poor TPT completion, whereas older age (≥25 years versus 15-24 years) was associated with higher TPT completion.

**Conclusion:** The targeted 100-day campaign dramatically increased TPT uptake and completion among PLHIV suggesting a viable catch up strategy to meet WHO guidelines.

## Introduction

Tuberculosis (TB) is the most frequent cause of Acquired Immunodeficiency Syndrome (AIDS)-related deaths worldwide despite the wide availability of antiretroviral therapy (ART)(1). Tuberculosis preventive treatment (TPT) reduces the risk of developing active TB(2) and TB-associated mortality among people living with Human Immunodeficiency Virus (PLHIV)(3, 4). Therefore, the World Health Organization (WHO) recommends incorporating TPT into routine HIV care for all adults, adolescents and children living with HIV without symptoms suggestive of TB or without active TB(5). Data from clinical trials, observational studies and routine care programs have demonstrated reduced TB incidence due to TPT use(6-9), with a dramatic drop in TB incidence in PLHIV when TPT is administered concomitantly with ART(6, 9, 10). However, TPT implementation has been slow, and compounded by suboptimal treatment completion rates (60 – 80%) in resource-limited setting countries (RLS)(4, 11, 12). In the recent 2020 guidelines for TB treatment, the WHO recommended shorter rifamycin-containing TPT regimens, which are associated with better completion rates(5). However, such regimens are not yet widely available and isoniazid monotherapy for 6 months is still the most frequently used regimen in RLS.

Uganda is among 30 high TB/HIV burden countries which contribute about 60% of the total TB/HIV burden globally. In 2017, about 40% of the new TB patients were HIV-positive according to the WHO TB Global report 2018 (13). However, in 2019, the proportion of TB notified cases among those who were newly enrolled into HIV care in 2019 was 6.7% (1). The Uganda Ministry of Health (MoH) recommends that all PLHIV with no symptoms suggestive of active TB disease are eligible for TPT regardless of CD4 count, ART status, history of TB treatment, and pregnancy status (14, 15). Before 2019, TPT uptake among PLHIV in Uganda was estimated at only about 16% to 17% (16, 17), with only 50%-60% of patients completing treatment(17, 18). Contributing factors to poor TPT uptake in Uganda included: inadequate TPT supply in health facilities, frequent drug stock-outs, poor patient adherence, limited TPT knowledge by health workers, lack of confidence in symptom-based TB screening alone, and fear of isoniazid resistance(11, 17). One of the interventions done by the Uganda MoH was the 100-da accelerated isoniazid preventive therapy (IPT) scale-up campaign launched on 3^rd^ July 2019(16). This campaign aimed to enroll 300,000 PLHIV on isoniazid preventive therapy at 1947 ART sites by 30th September 2019. There is scanty published data showing the trends of prevalence of TPT uptake and completion, and the impact of the 2019 Ugandan MoH 100-day TPT scale-up campaign on these trends.

This study aimed to examine the effect of the 100-days campaign on trends of TPT uptake and completion from routinely collected data from 2016 to 2019 at six public health facilities in Uganda. The study also explored patients’ characteristics associated with TPT uptake and TPT completion among PLHIV in care.

## Methods

### Setting, study design and population

This was a cross-sectional review of patients’ medical records at six HIV clinics in Kampala Capital City Authority (KCCA) public primary health care facilities (dubbed as health centers)(19), which are supported by the Infectious Diseases Institute (IDI), Uganda: Of these, five were health center level III (Kiswa, Kawaala, Komamboga, Kitebi, Kisugu) and one was a health center IV (Kisenyi). These are government health facilities providing free health care to about 2% of the population in Kampala district which approximated at 3.1 million in 2019(20, 21), with 6.9% HIV prevalence among adults aged 15-64 year during 2016-2017(22). The bacteriologically confirmed TB prevalence was estimated at about 401 per 100,000 in urban areas during 2014-2015(23).

During the study period (January 2016 and December 2019), isoniazid monotherapy for 6 months was the only TPT option available, and was administered according to the 2014 Uganda national guidelines(14), which indicate that all adolescents and adult PLHIV who are unlikely to have active TB should receive at least a six-months course of isoniazid as TPT, regardless of immune status, ART status, history of TB treatment, and pregnancy status. Vitamin B6 (pyridoxine) was concomitantly given with isoniazid to prevent occurrence of peripheral neuropathy (14).

### Inclusion and exclusion criteria

The study included all asymptomatic PLHIV aged 15 years and above, who were in care and visited the adult HIV clinics at least once each year from January 2016 to December 2019 at the six public health facilities, who met the eligibility criteria according to the national guidelines (14).

### Treatment procedure

According to the guidelines(14), patients were reviewed two weeks after initiating TPT to assess for severe side effects and reinforce adherence, and then received monthly refills and follow-up until 6 months after initiation. TPT was stopped if a patient was diagnosed with TB or had severe side effects: jaundice, fever, severe tingling and burning sensation, blurred vision, loss of vision, convulsions, or unusual bleeding. Information on TPT was recorded in the paper-based registers at each health facility, whereas HIV care data were recorded in an electronic medical records (EMR) database system. During the study period, ART was provided according to the 2016 Uganda consolidated guidelines for prevention and treatment of HIV in Uganda(24).

### Evaluation of TPT uptake and outcomes

TPT initiation and treatment outcome information was extracted from paper-based TPT registers at each of the six healthy facility studied. Because data was retrospectively collected, we additionally extracted prescribed drugs recorded in EMR for additional individuals for whom TPT was prescribed during the study period in order to reduce on underreporting due to possibility of missing information

### Data extraction and collection

A structured questionnaire for extraction of TPT information was designed and programmed into Open Data Kit (ODK)(25), a mobile data collection application. Data quality was controlled by applying data validation checks in the ODK database system, training of research assistants who extracted data from TPT registers and continuous review of data captured into ODK. Data from different registers (data sources) was linked using the patient HIV clinic identifiers. If the identifier was missing, then patient characteristics including; names, age, sex, home address, ART start date were used if available. To maintain anonymity of patients, papers that were used to record patients identifying information for linking data were destroyed immediately before leaving the health facility on day of data collection. For the HIV care information, a list of variables was created to guide the process of data extraction from the Uganda EMR database version 3.2.0 at each of the six health facilities studied. The data collection tools used are presented in the appendix. Data were collected from July 2020 to March 2021. Permission to access patients’ medical records in clinics studied was sought from the Directorate of Public Health and Environment, KCCA, Uganda.

### Statistical analysis

Data were analyzed using STATA software version 16.1 Special Edition (StataCorp, College Station, Texas, USA).

We analyzed TPT uptake and completion in each of the 4 years (2016-2019) to establish the trends. We examined change in TPT trends after the 100-days TPT accelerated campaigns in 2019. Baseline patient characteristics (listed in appendix) were defined at beginning of each year for the 4 years analyzed. For the analysis of TPT completion, baseline patient characteristics were defined as those at TPT initiation.

We used descriptive statistics (frequencies and percentages) to describe study participants across the four years studied. The prevalence of TPT uptake was estimated as the proportion of patients initiated on TPT among those who were eligible for TPT in a specific year. TPT completion was estimated as proportion of patients completed TPT among those initiated on TPT by year. Patients who were either lost-to-follow-up, died or stopped TPT were classified as non TPT completers. Factors associated with TPT uptake were examined by fitting Poisson regression with robust standard errors using generalized estimating equations (GEE) with exchangeable correlation structure to account for clustering of participants in the 6 clinics and repeated observation of participants over the 4 years. When the prevalence of the outcome is >10%, the odds ratio exaggerates the association, and is not approximately equal to the prevalence ratio (PR)(26).

Factors associated with TPT completion were examined by fitting Poisson regression model with cluster-correlated robust estimates of standard errors because the outcome was common (>10%). A separate multivariable modified Poisson regression model was fitted for females to examine the association between pregnancy status and TPT completion. All multivariable analyses were adjusted for the predefined baseline factors (gender, age groups, WHO stage, BMI categories, TB treatment history, pregnancy status at TPT initiation). Statistical significance testing was based on a 2-tailed Wald test and 5% significance level.

To examine difference in TPT uptake and completion across sub-groups of predefined baseline characteristics listed above, stratified regression models were fitted by entering interactions between the variable for calendar years and each of the predefined baseline covariates, in separate adjusted models.

We used variance inflation factors (VIFs) to evaluate multicollinearity in fitted models, and effect of missing data was assessed using the multiple imputation chained-equation (MICE) approach.

### Ethics consideration

The AIDS Support Organization Research Ethics Committee (TASOREC), Kampala, Uganda (number: TASOREC/085/19-UG-REC-009) and the Uganda National Council for Science and Technology, Kampala, Uganda (UNCST number: HS729ES) granted ethical approval for the study. Due to the retrospective study design and that this was a public health surveillance, the need for patient consent was waived.

## RESULTS

### Description of participants

Figure 1 shows the total numbers of patients who reported for HIV care at the six clinics by year; 95% of patients were eligible for TPT during each year and were included in the analysis of TPT uptake. Analysis of IPT completion was performed on 10,131 patients who had data in both paper-based TPT registers and the HIV care clinic electronic database. Data were missing on 10% and 4% patients for TPT uptake and treatment completion analyses, respectively. Across the 4 years, more than 70% were females (range: 73.5% -74.6%) and median age 34 years and inter-quartile range 28 to 41 years on overall over the 4 years. About 15% of the females were pregnant (range: 14.6 -16.0%). Almost all PLHIV were on ART due to test-and-treat expanded policy that was rolled out in 2016 in Uganda. However, because of the opt-out (24) option and that test and treat was not yet fully implemented in early 2016, some patients were not on ART but the proportion not on ART declined overtime from 9% in 2016 to about 2% in 2019. All collected baseline characteristics are shown in Table 1.

**Table 1:**
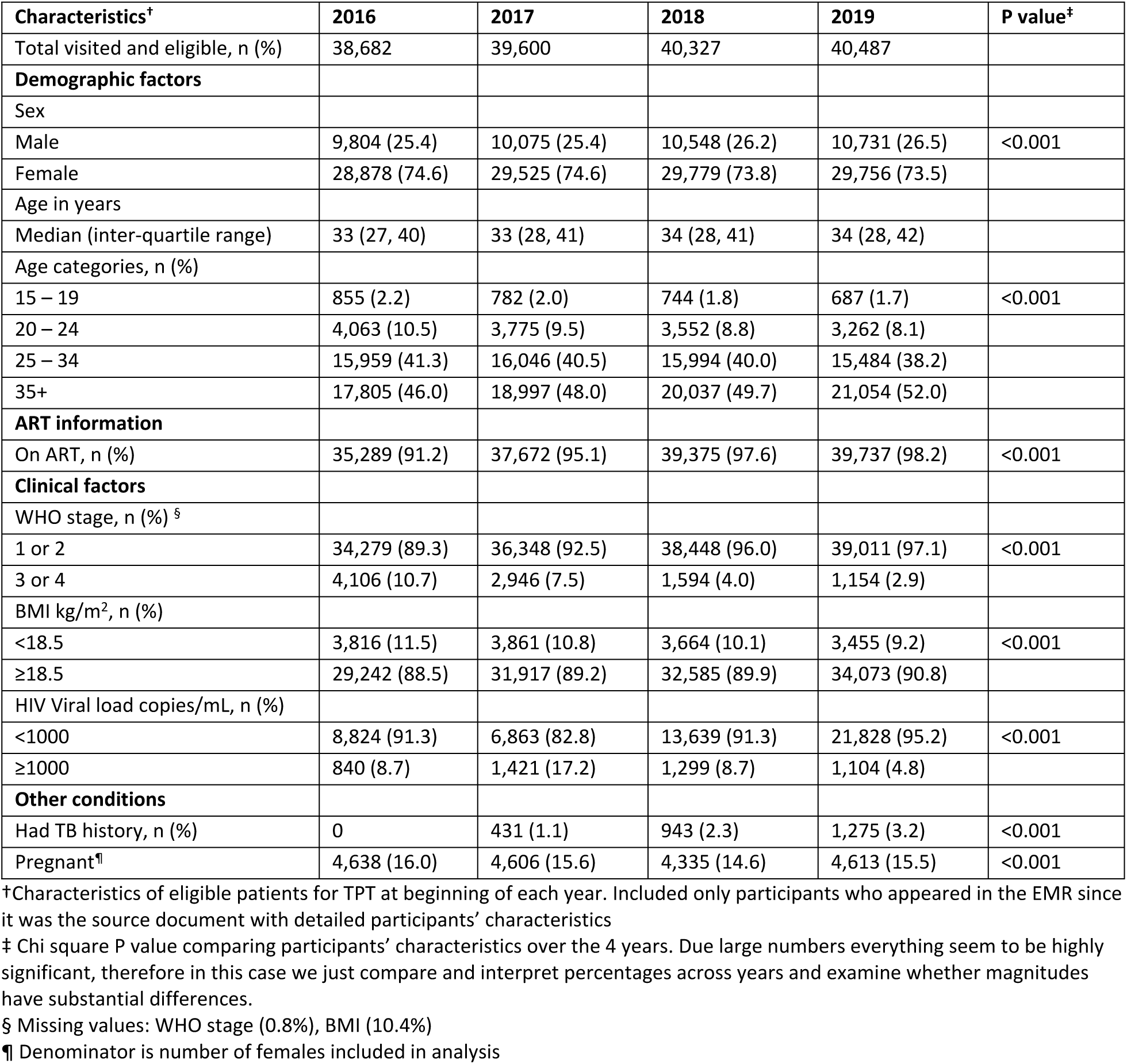
Patients’ baseline characteristics at TPT uptake by year.

| Characteristics<sup>†</sup> | 2016 | 2017 | 2018 | 2019 | P value<sup>‡</sup> |
| --- | --- | --- | --- | --- | --- |
| Total visited and eligible, n (%) | 38,682 | 39,600 | 40,327 | 40,487 |  |
| **Demographic factors** |  |  |  |  |  |
| Sex |  |  |  |  |  |
| Male | 9,804 (25.4) | 10,075 (25.4) | 10,548 (26.2) | 10,731 (26.5) | <0.001 |
| Female | 28,878 (74.6) | 29,525 (74.6) | 29,779 (73.8) | 29,756 (73.5) |  |
| Age in years |  |  |  |  |  |
| Median (inter-quartile range) | 33 (27, 40) | 33 (28, 41) | 34 (28, 41) | 34 (28, 42) |  |
| Age categories, n (%) |  |  |  |  |  |
| 15 – 19 | 855 (2.2) | 782 (2.0) | 744 (1.8) | 687 (1.7) | <0.001 |
| 20 – 24 | 4,063 (10.5) | 3,775 (9.5) | 3,552 (8.8) | 3,262 (8.1) |  |
| 25 – 34 | 15,959 (41.3) | 16,046 (40.5) | 15,994 (40.0) | 15,484 (38.2) |  |
| 35+ | 17,805 (46.0) | 18,997 (48.0) | 20,037 (49.7) | 21,054 (52.0) |  |
| **ART information** |  |  |  |  |  |
| On ART, n (%) | 35,289 (91.2) | 37,672 (95.1) | 39,375 (97.6) | 39,737 (98.2) | <0.001 |
| **Clinical factors** |  |  |  |  |  |
| WHO stage, n (%) <sup>§</sup> |  |  |  |  |  |
| 1 or 2 | 34,279 (89.3) | 36,348 (92.5) | 38,448 (96.0) | 39,011 (97.1) | <0.001 |
| 3 or 4 | 4,106 (10.7) | 2,946 (7.5) | 1,594 (4.0) | 1,154 (2.9) |  |
| BMI kg/m<sup>2</sup>, n (%) |  |  |  |  |  |
| <18.5 | 3,816 (11.5) | 3,861 (10.8) | 3,664 (10.1) | 3,455 (9.2) | <0.001 |
| ≥18.5 | 29,242 (88.5) | 31,917 (89.2) | 32,585 (89.9) | 34,073 (90.8) |  |
| HIV Viral load copies/mL, n (%) |  |  |  |  |  |
| <1000 | 8,824 (91.3) | 6,863 (82.8) | 13,639 (91.3) | 21,828 (95.2) | <0.001 |
| ≥1000 | 840 (8.7) | 1,421 (17.2) | 1,299 (8.7) | 1,104 (4.8) |  |
| **Other conditions** |  |  |  |  |  |
| Had TB history, n (%) | 0 | 431 (1.1) | 943 (2.3) | 1,275 (3.2) | <0.001 |
| Pregnant<sup>¶</sup> | 4,638 (16.0) | 4,606 (15.6) | 4,335 (14.6) | 4,613 (15.5) | <0.001 |

<sup>†</sup>Characteristics of eligible patients for TPT at beginning of each year. Included only participants who appeared in the EMR since it was the source document with detailed participants' characteristics
<sup>‡</sup> Chi square P value comparing participants' characteristics over the 4 years. Due large numbers everything seem to be highly significant, therefore in this case we just compare and interpret percentages across years and examine whether magnitudes have substantial differences.
<sup>§</sup> Missing values: WHO stage (0.8%), BMI (10.4%)
<sup>¶</sup> Denominator is number of females included in analysis

**Figure 1:**
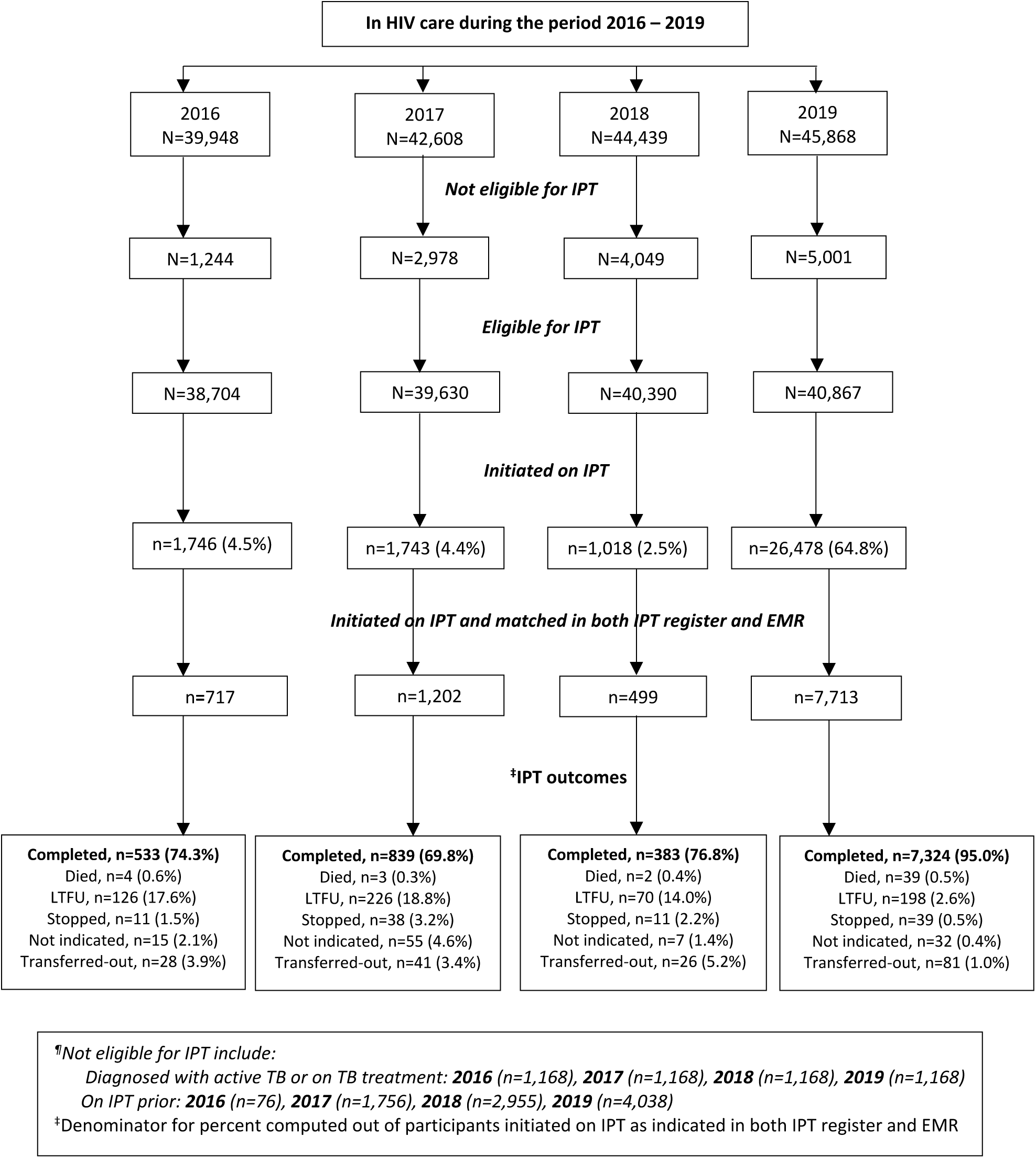
Patients’ flow diagram by the 4 years studied at the 6 health

### Trends of prevalence of TPT uptake

Among the eligible patients who had a clinic encounter at the selected health facilities during the period studied, there was very low TPT uptake with a declining trend from 2016 to 2018 (prevalence of TPT uptake 4.5%, 4.4% and 2.5% during 2016, 2017 and 2018 respectively). However, as expected in 2019 uptake increased dramatically by about 25% (relative increase) from 2.5% in 2018 to 64.8% in 2019 (Figure 1). At multivariable analysis using modified Poisson GEE model, prevalence of TPT uptake was 14.7 times higher in 2019 compared to that in 2016 (adjusted Prevalence Ratio [aPR] = 14.7, 95% confidence interval [95%CI] = 14.1, 15.4, and P value<0.001) and it was 26.0 times higher compared to 2018 (aPR = 26.0 [95%CI 24.5, 27.7], and P value<0.001). The analysis was adjusted for participant baseline characteristics at beginning of each year (sex, age categories, ART status, WHO stage, BMI categories, TB history) (Table 2).

**Table 2:** Factors associated with TPT uptake.

| Factors | Crude PR (95%CI) † | P-value | Adjusted PR (95%CI) † | P-value |
| --- | --- | --- | --- | --- |
| **Calendar year\*** |  |  |  |  |
| 2016 | 1 |  | 1 |  |
| 2017 | 0.96 (0.90, 1.02) | 0.185 | 0.98 (0.92,1.04) | 0.530 |
| 2018 | 0.54 (0.50, 0.58) | <0.001 | 0.56 (0.52,0.61) | <0.001 |
| 2019<sup>‡</sup> | 13.9 (13.3, 14.6) | <0.001 | 14.7(14.1,15.4) | <0.001 |
| **Demographic factors** |  |  |  |  |
| Sex |  |  |  |  |
| Male | 1 |  | 1 |  |
| Female | 0.99 (0.97, 1.01) | 0.278 | 0.96 (0.94, 0.98) | <0.001 |
| Age in years, n (%) |  |  |  |  |
| 15 – 19 | 1 |  | 1 |  |
| 20 – 24 | 0.86 (0.81, 0.93) | <0.001 | 0.88 (0.83, 0.93) | <0.001 |
| 25 – 34 | 0.76 (0.72, 0.81) | <0.001 | 0.76 (0.72, 0.81) | <0.001 |
| 35+ | 0.70 (0.65, 0.74) | <0.001 | 0.67 (0.63, 0.71) | <0.001 |
| **Clinical factors** |  |  |  |  |
| On ART<sup>§</sup> |  |  |  |  |
| No | 1 |  | 1 |  |
| Yes | 0.96 (0.91, 1.01) | 0.139 | 0.51 (0.48, 0.55) | <0.001 |
| WHO stage<sup>§</sup> |  |  |  |  |
| 1 or 2 | 1 |  | 1 |  |
| 3 or 4 | 0.58 (0.55, 0.61) | <0.001 | 0.92 (0.87, 0.97) | 0.001 |
| BMI kg/m<sup>2</sup> § |  |  |  |  |
| <18.5 | 1 |  | 1 |  |
| ≥18.5 | 1.10 (1.06, 1.13) | <0.001 | 1.03 (1.00, 1.06) | 0.020 |
| TB treatment history<sup>§</sup> |  |  |  |  |
| No | 1 |  | 1 |  |
| Yes | 1.54 (1.46, 1.62) | <0.001 | 0.90 (0.86, 0.94) | <0.001 |

\* Number of completed case analyzed (Year2016 n= 33,021, Year2017 n=35,718, Year2018 n=36,198, Year2019 n=37,418). Proportion of missing data on at-least one of the variables per year in adjusted model, (Year2016 [15%], Year2017 [10%], Year2018 [10%], Year2019 [8%]).
† **PR** –Prevalence Ratio estimated using Generalized Estimating Equations (GEE) with modified Poisson regression model and robust standard errors accounting for repeated observation of participants over the 4 years CI – confidence interval
♀ adjusted prevalence ratio comparing TPT uptake in 2019 and 2018 (aPR [95%CI] = 26.0 [24.5, 27.7], P value<0.001)
† Refitting the same model for women only to examine TPT uptake by pregnancy status, adjusted Prevalence Ratio for TPT uptake = 1.25 (95%CI = 1.22, 1.28), P value <0.001, among pregnant compared non-pregnant (reference category)

Stratified analysis indicated: TPT uptake was similar across age groups and sex of participants over the 4 years studied (Figures 2a and 2b). TPT uptake was significantly higher among patients who were not on ART compared to those who were on ART before 2018 (predicted mean probabilities of TPT were 27.4% vs 3.2% in 2016, 13.5% vs 4.3% in 2017, 1.0% vs 2.7% in 2018, 54.9% vs 69.3% in 2019 respectively) (Figure 3a). Also, TPT uptake was slightly higher among pregnant women compared to those not pregnant (predicted probabilities were 4.5% vs 4.6% in 2016, 3.9% vs 9.5% in 2017, 2.3 % vs 4.0 % in 2018, 68.9 % vs 81.3 % in 2019) (Figure 3b). Nonetheless, in 2019 TPT uptake increased uniformly in all categories.

**Figure 2a:**
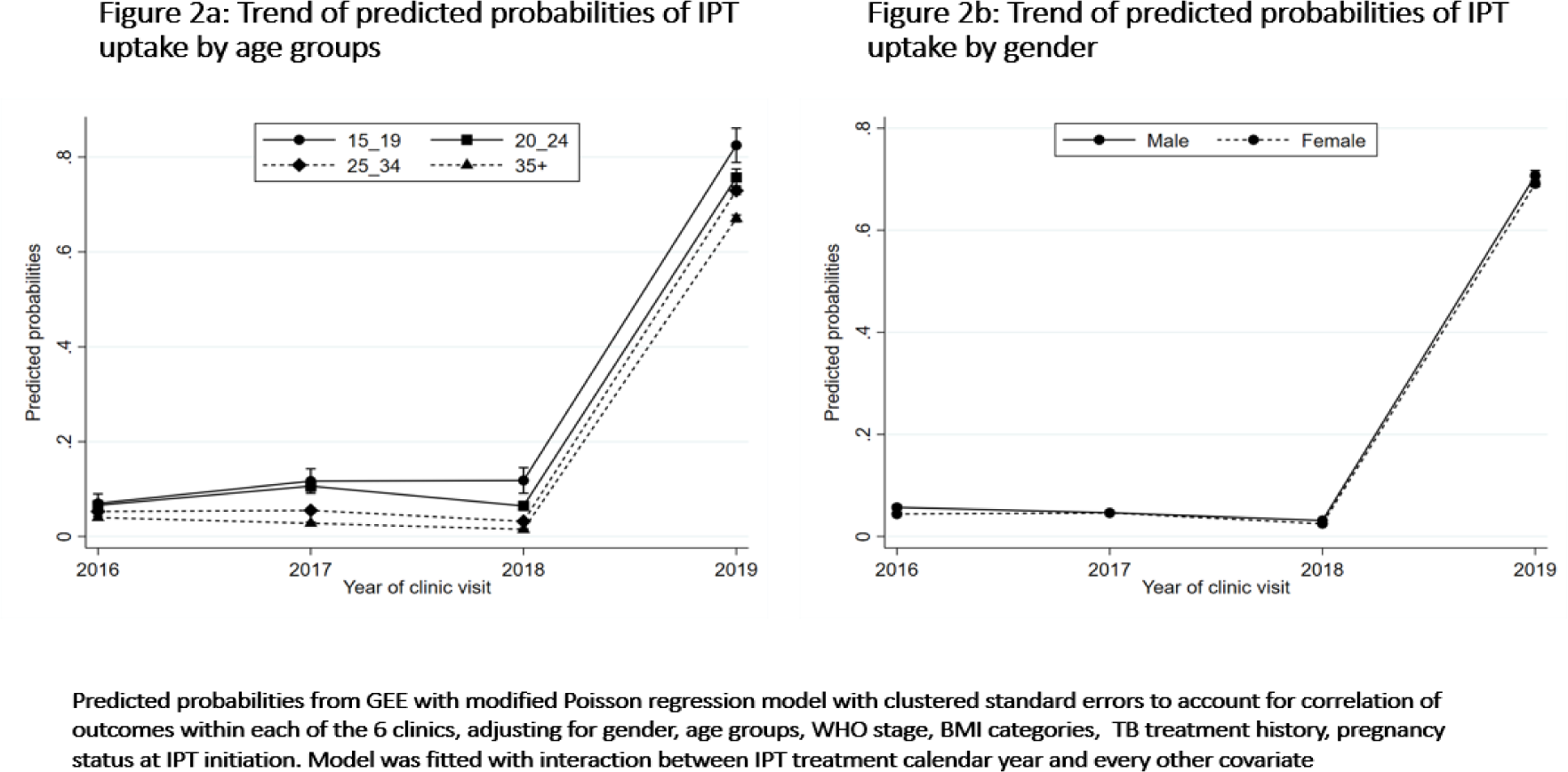
Trend of predicted probabilities of TPT update by age groups Figure 2b: Trend of predicted probabilities of TPT update by gender

**Figure 3a:**
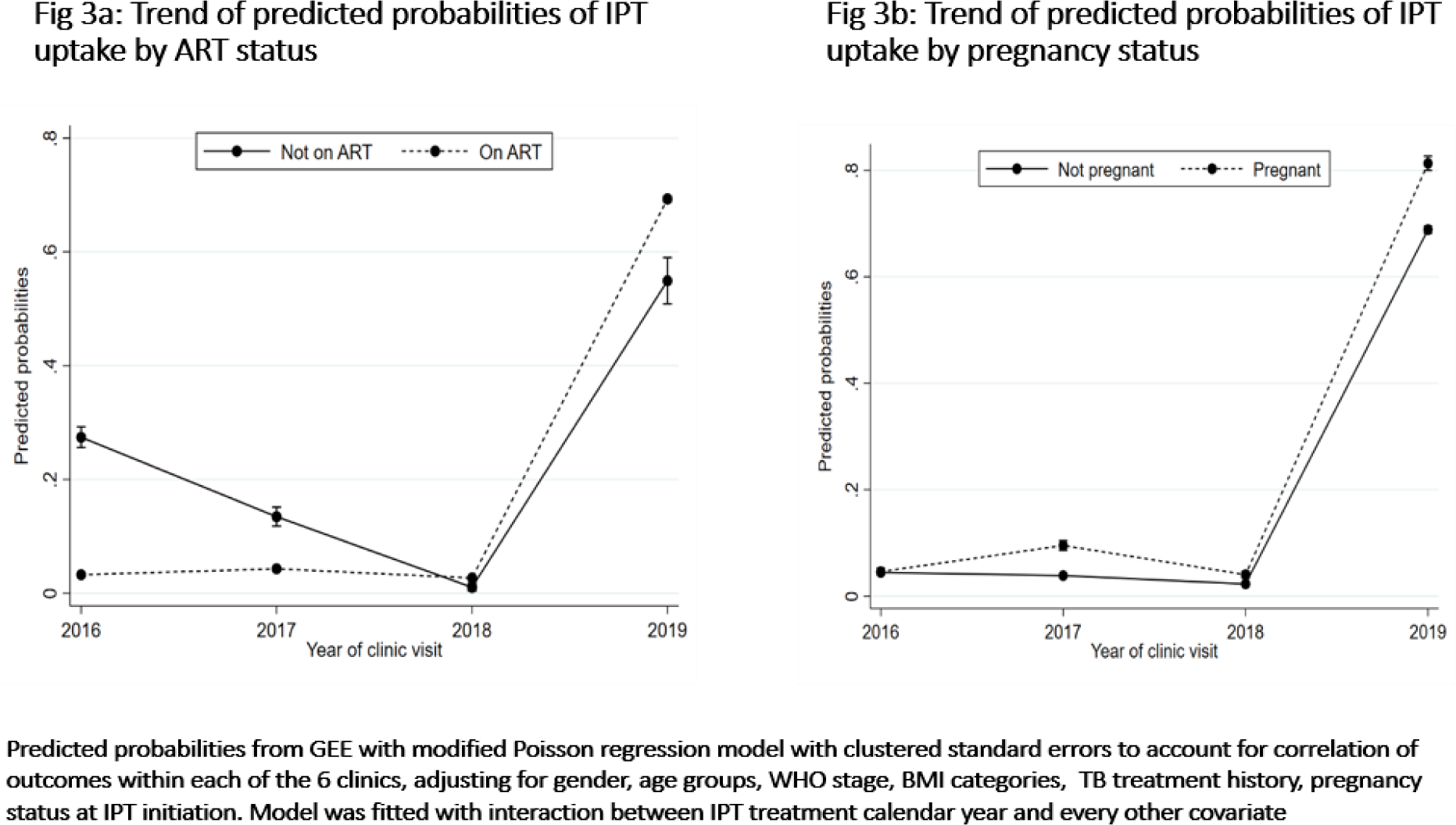
Trend of predicted probabilities of TPT update by ART status Figure 3b: Trend of predicted probabilities of TPT update by pregnancy status

### Trends of TPT completion

TPT completion was consistently high at about 80% between years 2016 – 2018 but increased by 17% (relative increase) in 2019 compared to that in 2018 (TPT completion was 81.2%, 76.3%, 82.6% and 96.5% in year 2016, 2017, 2018 and 2019 respectively). The increase in TPT completion between 2018 and 2019 remained significant even after adjusting for patients’ baseline characteristics (aPR [95%CI] = 1.12 [1.04, 1.21], P value=0.003). TPT completion was significantly higher among patients aged 25 years and above compared to those below 25 years (aPR [95%CI] = 1.06 [0.98, 1.14], 1.15 [1.05, 1.25], 1.18 [1.08, 1.30] for ages 20 – 24, 25 – 34 and 35+years respectively with 15 – 19 years as reference age group). Patients not on ART or newly initiated on ART at TPT start were significantly associated with lower TPT completion compared to the ART experienced (aPR [95%CI] = 0.80 [0.73, 0.89], 0.90 [0.86, 0.95], respectively). There was no sufficient evidence for the association of TPT completion with sex (Wald P value = 0.156), WHO stage (Wald P value = 0.240), BMI (Wald P value = 0.844) and pregnancy status (Wald P value = 0.104) (table 3). There was no substantial change in estimates (both prevalence ratios and 95%CI) after refitting the model after multiple imputation of missing values.

**Table 3:** TPT completion and associated factors.

| Factor | Total* | IPT completion, n (%) | Crude PR (95%CI) † | P-value | Adjusted PR (95%CI) † | P-value |
| --- | --- | --- | --- | --- | --- | --- |
| Calendar year |  |  |  |  |  |  |
| 2016 | 629 | 511 (81.2) | 1 |  | 1 |  |
| 2017 | 1,044 | 797 (76.3) | 0.93 (0.81, 1.09) | 0.403 | 0.89 (0.76, 1.06) | 0.184 |
| 2018 | 442 | 365 (82.6) | 1.02 (0.87, 1.19) | 0.839 | 0.97 (0.85, 1.11) | 0.711 |
| 2019<sup>‡</sup> | 7,307 | 7,048 (96.5) | 1.19 (1.04, 1.36) | 0.012 | 1.09 (0.97, 1.22)<sup>‡</sup> | 0.138 |
| **Demographic factors** |  |  |  |  |  |  |
| Sex |  |  |  |  |  |  |
| Male | 2,619 | 2,452 (93.6) | 1 |  | 1 |  |
| Female | 6,803 | 6,269 (92.2) | 0.98 (0.97, 0.99) | 0.039 | 0.99 (0.97, 1.00) | 0.155 |
| Age in years, n (%) |  |  |  |  |  |  |
| 15 – 19 | 198 | 158 (79.8) | 1 |  | 1 |  |
| 20 – 24 | 917 | 767 (83.6) | 1.05 (0.91, 1.21) | 0.513 | 1.06 (0.98, 1.14) | 0.150 |
| 25 – 34 | 3,829 | 3,493 (91.2) | 1.14 (0.99, 1.33) | 0.077 | 1.15 (1.05, 1.25) | 0.002 |
| 35+ | 4,478 | 4,303 (96.1) | 1.20 (1.05, 1.38) | 0.008 | 1.18 (1.08, 1.30) | <0.001 |
| **Clinical factors** |  |  |  |  |  |  |
| ART status |  |  |  |  |  |  |
| Not on ART | 314 | 237 (75.5) | 0.78 (0.68, 0.89) | <0.001 | 0.80 (0.73, 0.89) | <0.001 |
| Newly on ART<sup>†</sup> | 2,565 | 2,124 (82.8) | 0.85 (0.80, 0.91) | <0.001 | 0.90 (0.86, 0.95) | <0.001 |
| ART experienced | 6,543 | 6,360 (97.2) | 1 |  | 1 |  |
| WHO stage<sup>§</sup> |  |  |  |  |  |  |
| 1 or 2 | 9,160 | 8,491 (92.7) | 1 |  | 1 |  |
| 3 or 4 | 262 | 230 (87.8) | 0.95 (0.93, 0.97) | <0.001 | 0.97 (0.93, 1.02) | 0.240 |
| BMI kg/m<sup>2</sup> <sup>§</sup> |  |  |  |  |  |  |
| <18.5 | 877 | 797 (90.9) | 1 |  | 1 |  |
| ≥18.5 | 8,545 | 7,924 (92.7) | 1.02 (0.99, 1.05) | 0.100 | 1.00 (0.98, 1.03) | 0.843 |
| TB history |  |  |  |  |  |  |
| No | 9,191 | 8,499 (92.5) | 1 |  | 1 |  |
| Yes | 231 | 222 (96.1) | 1.04 (1.01, 1.07) | 0.011 | 0.98 (0.97, 0.99) | 0.009 |

\* Number of completed case analyzed, N= 9,682/10,131 (96%). Missing on at-least one of the variables in adjusted model, n=449 (4%).
<sup>†</sup> PR – Prevalence Ratio estimated using modified Poisson regression model with cluster standard errors to account for clustering since data was collected from different clinics.
CI – confidence interval, BMI – Body Mass Index
♀ adjusted prevalence ratio comparing IPT completion in 2019 and 2018 (aPR [95%CI] = 1.12 [1.04, 1.21], P value=0.003)
§ Missing values: outcome (n=285, 3%), WHO stage (n=98, 1%), BMI (n=396, 4%)
‡ Newly on ART includes participants who were on ART for ≤ 3 months at TPT initiation.
Refitting the same model for women only to examine TPT uptake by pregnancy status, adjusted Prevalence Ratio for IPT completion = 0.96 (95%CI = 0.92, 1.01), P value=0.104, among pregnant compared non-pregnant (reference category)

At stratified analysis, compared to the ART experienced, TPT completion was slightly lower before 2019 compared to that in 2019 among those not on ART (predicted mean probabilities of IPT uptake : 73.1% vs 87.4%, 53.1% vs 87.2%, 75.5% vs 96.8%, 97.0% vs 97.1% during 2016, 2017, 2018, 2019 respectively), and among newly initiated on ART (predicted mean probabilities of IPT uptake : 84.8% vs 87.4%, 77.5% vs 87.2%, 81.0% vs 96.8%, 90.0% vs 97.1% during 2016, 2017, 2018, 2019 respectively) (Figure 5a). Similarly, TPT completion was lower before 2019 among patients aged 19 years or below (Figure 4a) and among pregnant females (Figure 5b). In 2019 IPT completion uniformly increased across all groups.

**Figure 4a:**
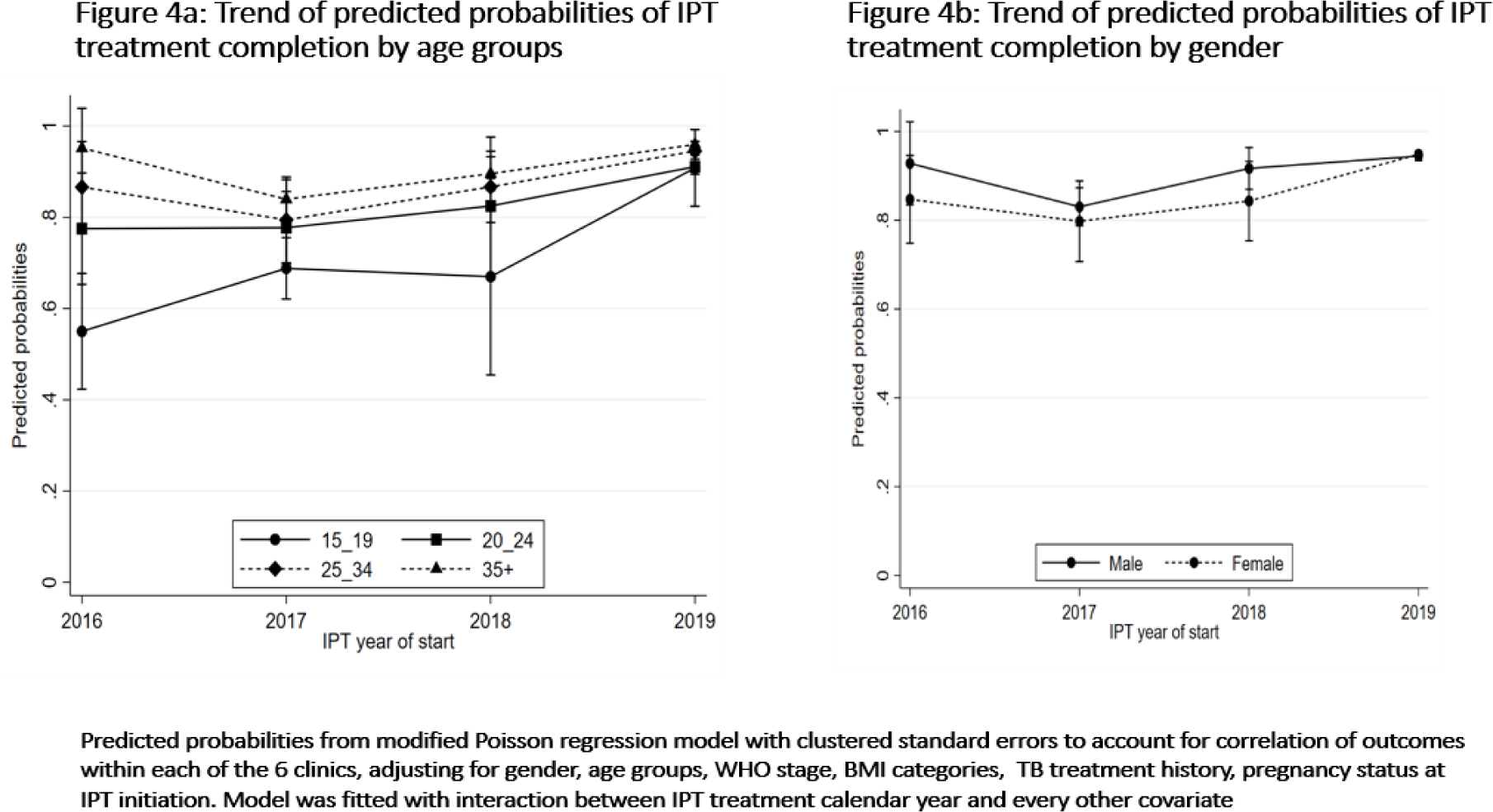
Trend of predicted probabilities of IPT completion by age groups Figure 4b: Trend of predicted probabilities of IPT completion by gender

**Figure 5a:**
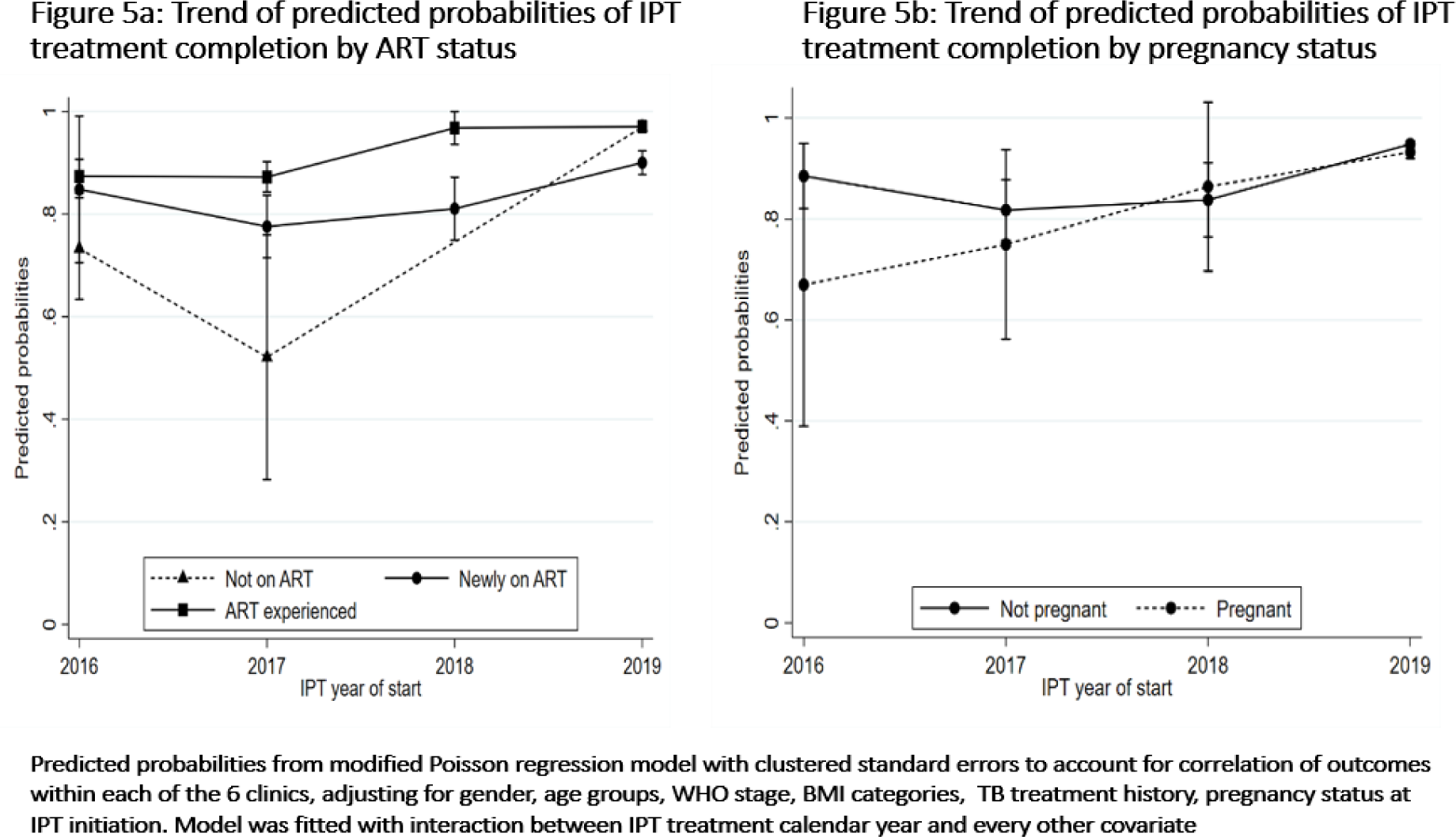
Trend of predicted probabilities of IPT completion by ART status Figure 5b: Trend of predicted probabilities of IPT completion by pregnancy status

For the IPT non-completers, the proportions of those who died was consistently below 1% (ranged: 0.3% to 0.6%), whereas proportion of LTFU dropped systematically by 85% (relative drop) from 17.6% (126/717) in 2016 to 2.6% (198/7,713) in 2019 (figure 1). Participants aged <25years, not on ART or newly on ART and those pregnant during TPT had higher proportions of LTFUs compared to their counterparts (table A1, appendix1).

## Discussion

The 100 day campaign was a major intervention undertaken by the Ugandan Ministry of Health (MoH) in 2019 and increased IPT uptake overall. Despite the large number initiated, the TPT completion rates also increased substantially. This was achieved using existing health care resources and system strengthening (16). The campaign objectives included enrolling 300,000 PLHIV on IPT in 100 days in order to improve IPT coverage from about 30% before the campaign July 2019) to at least 50% of all the PHIV in care by the end of the campaign (October 2019) and ensure 100% IPT completion for all enrolled patients before and after the campaign. Based on these results, we can conclude that the 100 day TPT campaign in Uganda was highly successful. Given that many resource-limited settings are still struggling to achieve high rates of TPT coverage among the PLHIV, this is an intervention that we can recommend in such settings to improve TPT programing.

TPT uptake was higher among newly enrolled PLHIV in care not on ART and among pregnant women living with HIV before 2018. The national guidance before 2018 was to enroll only the newly enrolled PLHIV in care; however, in 2019, there was an expanded eligibility for TPT enrollment to all PLHIV who had never taken a course of TPT before (16). With the expanded TPT enrollment eligibility, increased TPT availability during the 100 day campaign in 2019(16), we observed improved uptake across all patient groups. TPT uptake is however still suboptimal on overall, and still below 100% the desired level. Previously, low TPT uptake had been reported in another prospective study conducted in 2017 in Uganda by Kalema et.al (17) where of 372 PLHIV participants who were eligible, only 17% initiated TPT. Although it was impossible to explore reasons for poor TPT uptake given the retrospective nature of our study, some of the reasons cited for low TPT uptake highlighted in Kalema et.al study included: limited capacity of clinicians to exclude TB using symptoms alone, fear of promoting drug resistance due to isoniazid monotherapy and inconsistent TPT drug supplies(17).

In prospective research studies, many countries in sub-Saharan Africa (SSA) have shown similar success with TPT completion rates above 80%(7, 27, 28). In this report we show high completion rates of TPT in a program setting which can be attributed to, good coordination between the AIDS Control and National TB/Leprosy Programs, consistent supply of INH and vitamin B6, substantial support from partners like U.S President’s Emergency Plan for AIDS Relief (PEPFAR), United States Agency for International Development (USAID), Centers for Disease Control (CDC), among other, partner engagement, facility mentorship activities and improved reporting (16).

We observed that patients not on ART or newly initiated on ART were associated with low IPT completion before 2019 compared to their ART experienced counterparts. Similarly, some previous studies found that TPT completion was associated with being on ART (7, 29). Patients who are new or not on ART are likely to have stigma (30, 31), poor adherence (32), and lack of understanding of the role of TB prevention in the absence of symptoms (33).

The higher TPT completion among older persons (aged ≥25 years) compared to that among younger persons (aged 15 – 24 years) found in our study could be attributed to poor treatment adherence among young persons as cited in other previous studies (32). This indicates that there is need for better strategies focusing at improvement treatment adherence among young PLHIV in order to achieve better TPT completion rates.

## Limitations

In this retrospective review of routinely collected data, we had the following limitations: firstly, there is a possibility that we included some patients not eligible for IPT due to undocumented comorbidities. This could have exaggerated the denominator leading to under-estimation of IPT uptake. Secondly, at analysis we only included patients whose clinical identifiers matched in both HIV care data and IPT data. However, the baseline characteristics of patients included compared to those excluded were similar, implying little potential effect of selection bias. Lastly, we could not report on TPT side effects because this information was largely missing in the data sources. Results from our study can be generalized to other similar resource-limited settings with high of TB and HIV coinfections.

## Conclusion

The 100 day TPT campaign intervention in Uganda was highly successful in initiation and completion of TPT among PLHIV and overcame many of the barriers that have plagued scale up of TPT in national programs. From above findings we recommend more counselling sessions for pregnant women living with HIV, PLHIV below the age of 25 years and those newly enrolled on ART to ensure better TPT completion rates. Also, given that Uganda is rolling out rifapentine and isoniazid once weekly for 3 months (3HP) as per the national TPT guidelines (15), young people living with HIV should be prioritized for this regimen given its shorter duration, better completion rates and fewer adverse events (5). Also, there is need for constant refresher trainings for health workers to understand importance of TPT, and consistent TPT supply at health facilities.

## Data Availability

Data cannot be shared publicly because of confidentiality. Data are available on request from the Infectious Diseases Insitute, Institutional Data Access / Ethics Committee (contact via) for researchers who meet the criteria for access to confidential data.

## Acknowledgments

The program is supported by the Presidents Emergency Plan for AIDS Relief (PEPFAR) through the United States Centers for Disease Control (CDC) and Prevention (terms of Cooperative Agreement NU2GGH002022).

The authors acknowledge contributions of various staff from the Infectious Diseases Institute: Ms. Aidah Nanvuma, Mr. Grace Banturaki, Mr. Godwin Anguzu, Mr. Edison Katunguka; Research assistants: Mr. Daniel Kirumira, Ms. Jessica Masika, Ms. Carol Kaidu, Ms. Patricia Asiimwe, Mr. Martin Atugonza, Ms. Namada Moureen Juma; staff at the KCCA Directorate of Public Health and Environment office in Uganda: Dr. Daniel Okello, Ms. Phiona Nazziwa; and staff and in-charges at the 6 KCCA health care facilities who assisted in retrieval of patients’ records. We finally thank the CDC staff who reviewed the manuscript: Deus Lukoye, Grace Namayanja, Donna Kabatesi, Nelson Lisa J, and Ssempiira Julius.

## Disclaimer

The funders had no role in study design, data collection and analysis, decision to publish, or preparation of the manuscript.

## IPT uptake

**Stratified analysis: IPT uptake by subgroups (interactions between calendar years X patients characteristics)**

## IPT completion

**Stratified analysis: IPT completion by subgroups (interactions between calendar years X patients characteristics)**

### Appendix 1

#### Data variables

The following variables were extracted from the Uganda EMR database: Patients’ socio-demographics (age, birthdates, gender, marital status), clinical factors (clinic visit date, weight in kilograms, height in meters, ART status at IPT initiation, ART start date, WHO stage, HIV/RNA viral load (absolute), pregnancy status, TB treatment history, Isoniazid prophylaxis start date, and place of residence.

The following information were extracted from the TPT registers: IPT initiation date, IPT side effects, IPT outcome (completed 6-month IPT dose, loss to follow-up, died, stopped, transferred to another health facility), medical reasons for stopping IPT (including side effects, developed active TB).

### Appendix 2

**Table A1:** Detailed IPT outcomes by participants’ baseline characteristics at TPT start.

| Characteristics | Treatment outcomes |  |  |  |  |  | Total<br>n (%) | P value |
| --- | --- | --- | --- | --- | --- | --- | --- | --- |
|  | Completed<br>n (%) | Died<br>n (%) | LTFU<br>n (%) | Stopped<br>n (%) | Outcome not<br>indicated<br>n (%) | Transferred<br>out<br>n (%) |  |  |
| Year of IPT<br>treatment start |  |  |  |  |  |  |  |  |
| 2016 | 533 (74.3) | 4 (0.6) | 126 (17.6) | 11 (1.5) | 15 (2.1) | 28 (3.9) | 717 (100.0) | <0.001 |
| 2017 | 839 (69.8) | 3 (0.2) | 226 (18.8) | 38 (3.2) | 55 (4.6) | 41 (3.4) | 1202 (100.0) |  |
| 2018 | 383 (76.8) | 2 (0.4) | 70 (14.0) | 11 (2.2) | 7 (1.4) | 26 (5.2) | 499 (100.0) |  |
| 2019 | 7324 (95.0) | 39 (0.5) | 198 (2.6) | 39 (0.5) | 32 (0.4) | 81 (1.0) | 7713 (100.0) |  |
| Sex |  |  |  |  |  |  |  |  |
| Male | 2557 (91.5) | 15 (0.6) | 143 (5.1) | 23 (0.8) | 18 (0.6) | 38 (1.4) | 2794 (100.0) | 0.002 |
| Female | 6522 (88.9) | 33 (0.5) | 477 (6.5) | 76 (1.0) | 91 (1.2) | 138 (1.9) | 7337 (100.0) |  |
| Age in years at IPT<br>start |  |  |  |  |  |  |  |  |
| 15 – 24 | 982 (76.4) | 5 (0.4) | 181 (14.1) | 17 (1.3) | 41 (3.2) | 59 (4.6) | 1285 (100.0) | <0.001 |
| 25 – 34 | 3652 (87.8) | 25 (0.6) | 305 (7.3) | 43 (1.0) | 51 (1.2) | 85 (2.1) | 4161 (100.0) |  |
| ≥ 35 | 4445 (94.9) | 18 (0.4) | 134 (2.9) | 39 (0.8) | 17 (0.3) | 32 (0.7) | 4685 (100.0) |  |
| ART status at IPT |  |  |  |  |  |  |  |  |
| Not on ART | 244 (65.4) | 3 (0.8) | 90 (24.1) | 6 (1.6) | 6 (1.6) | 24 (6.5) | 373 (100.0) | <0.001 |
| Newly on ART<sup>†</sup> | 2290 (77.5) | 16 (0.6) | 406 (13.7) | 57 (1.9) | 72 (2.4) | 116 (3.9) | 2957 (100.0) |  |
| ART<br>experienced<sup>††</sup> | 6545 (96.3) | 29 (0.4) | 124 (1.8) | 36 (0.5) | 31 (0.5) | 36 (0.5) | 6801 (100.0) |  |
| WHO HIV stage |  |  |  |  |  |  |  |  |
| Stage 1 or 2 | 8767 (89.9) | 45 (0.5) | 581 (6.0) | 92 (0.9) | 104 (1.1) | 162 (1.7) | 9751 (100.0) | 0.014 |
| Stage 3 or 4 | 239 (84.8) | 3 (1.1) | 23 (8.2) | 7 (2.5) | 2 (0.7) | 8 (2.8) | 282 (100.0) |  |
| BMI at IPT start<br>(kgs/m<sup>2</sup>) |  |  |  |  |  |  |  |  |
| <18.5 | 801 (89.0) | 7 (0.8) | 58 (6.4) | 15 (1.7) | 7 (0.8) | 12 (1.3) | 900 (100.0) | 0.086 |
| ≥18.5 | 7961 (90.1) | 37 (0.4) | 514 (5.8) | 78 (0.9) | 100 (1.1) | 145 (1.6) | 8835 (100.0) |  |
| Pregnant during IPT |  |  |  |  |  |  |  |  |
| No | 2525 (91.1) | 26 (0.4) | 310 (5.4) | 57 (1.0) | 39 (0.7) | 80 (1.4) | 5764 (100.0) | <0.001 |
| Yes | 1270 (80.7) | 7 (0.5) | 167 (10.6) | 19 (1.2) | 52 (3.3) | 58 (3.7) | 1573 (100.0) |  |

<sup>†</sup> Newly on ART defined as being on ART for less than 3 months at IPT start
<sup>††</sup> ART experienced defined as being on ART for more than 3 months at IPT start

**Table A2.** Proportion of TPT uptake across participants characteristics by year (un adjusted)

| **Factors** | **2016**<br>**N=33,021** | **2017**<br>**N=35,718** | **2018**<br>**N=36,198** | **2019**<br>**N=37,418** |
| --- | --- | --- | --- | --- |
|  | % (n/N) | % (n/N) | % (n/N) | % (n/N) |
| **Demographic factors** |  |  |  |  |
| **Sex** |  |  |  |  |
| Male | 5.7 (475/8,358) | 4.4 (396/8,923) | 2.9 (268/9,315) | 67.1 (6,577/9,800) |
| Female | 4.5 (1,108/24,663) | 4.6 (1,236/26,795) | 2.4 (644/26,883) | 68.4 (18,913/27,618) |
| *P-value* | <0.001 | 0.493 | 0.011 | 0.013 |
| **Age in years** |  |  |  |  |
| 15 – 19 | 7.0 (39/558) | 11.8 (67/570) | 11.4 (63/551) | 79.9 (426/533) |
| 20 – 24 | 6.7 (193/2,863) | 10.5 (320/3036) | 6.2 (166/2,694) | 73.1 (1,956/2,676) |
| 25 – 34 | 5.3 (706/13,357) | 5.4 (767/14,342) | 3.0 (424/14,139) | 70.5 (10,001/14,181) |
| 35+ | 4.0 (645/16,243) | 2.7 (478/17,770) | 1.4 (259/18,814) | 65.4 (13,107/20,028) |
| *P-value* | <0.001 | <0.001 | <0.001 | <0.001 |
| **Clinical factors** |  |  |  |  |
| **On ART** |  |  |  |  |
| No | 27.9 (616/2,209) | 13.4 (206/1,533) | 0.9 (7/781) | 52.5 (322/613) |
| Yes | 3.1 (967/30,812) | 4.2 (1,426/34,185) | 2.6 (905/35,417) | 68.4 (25,168/36,805) |
| *P-value* | <0.001 | <0.001 | 0.003 | <0.001 |
| **WHO stage** |  |  |  |  |
| 1 or 2 | 4.8 (1,417/29,530) | 4.7 (1,553/33,074) | 2.5 (877/34,855) | 68.2 (24,867/36,442) |
| 3 or 4 | 4.8 (166/3,491) | 3.0 (79/2,644) | 2.6 (35/1,343) | 63.8 (623/976) |
| *P-value* | 0.910 | <0.001 | 0.836 | 0.004 |
| **BMI kg/m2** |  |  |  |  |
| <18.5 | 5.5 (211/3,811) | 4.2 (162/3,854) | 3.2 (118/3,650) | 65.6 (2,259/3,444) |
| ≥18.5 | 4.7 (1,372/29,210) | 4.6 (1,470/31,864) | 2.4 (794/32,548) | 68.4 (23,231/33,974) |
| *P-value* | 0.023 | 0.250 | 0.004 | 0.001 |
| **TB treatment history** |  |  |  |  |
| No | 4.8 (1,583/33,021) | 4.6 (1,629/35,314) | 2.6 (909/35,301) | 68.3 (24,714/36,187) |
| Yes | 0 | 0.7 (3/404) | 0.3 (3/897) | 63.0 (776/1,231) |
| *P-value* |  | <0.001 | <0.001 | <0.001 |

